# Opinion of Health Care Providers on Birth Companions in Obstetrics Department of a Tertiary Care Hospital in North India

**DOI:** 10.1101/2021.06.24.21259462

**Authors:** Tanvi Sarwal, Yamini Sarwal, Shakun Tyagi, Rakesh Sarwal

## Abstract

**Background:** Despite impressive improvements in institutional births and a fall in maternal mortality, satisfaction of women with birthing experience in public health institutions is low (68%). Birth Companion is an important part of the Labour room Quality Improvement (LaQshya) programme introduced by the Government of India in 2017. Despite mandates, implementation of the concept has been unsatisfactory (9%), even though the importance of Birth Companion has increased due to enhanced risk posed by COVID-19. Little is known about awareness among health care providers on Birth Companions, perceived barriers or their suggestions.

**Methods:** We canvassed a 15-question instrument using ordinal scales on 151 health care providers comprising consultants, post graduates, residents, and nurses (response rate 69%) in the department of Obstetrics & Gynecology, Lok Nayak Hospital, Delhi, India to gauge their awareness and opinions about Birth Companions.

**Results:** Most health care providers across all categories were aware of the concept (93%), World Health Organization’s recommendation (83%) and Government’s instructions for its hospitals (68%) that every woman should be accompanied by a Birth Companion of her choice during labour. Birth Companions of choice suggested by them were the mother (70%), husband (69%). sister (46%) or nurse (43%). Most health care providers agreed that a Birth Companion should wear clean clothes (95%), be free from any communicable disease (91%), stay with the pregnant woman throughout the process of labour (74%) and should have herself gone though labour (42%). Almost all providers (95%) agreed that the presence of a Birth Companion during labour will be beneficial, as they would provide emotional support (99%), boost the woman’s confidence (98%), provide comfort measures like massage (95%), early initiation of breastfeeding (93%), reduce post-partum depression (91%), humanize labour (83%), reduce need for analgesia (70%) and increase spontaneous vaginal births (69%). Yet support for its introduction in their hospital was low (59%). Staff nurses had reservations (62%) with only 40% of those who believed Birth Companion to be beneficial approving of its introduction in their hospital. Over-crowding in labour room and privacy concerns for other women were identified as key barriers.

**Conclusion:** Even though most health care providers were aware of and convinced of multiple benefits of Birth Companion during labour, lack of adequate infrastructure in the labour room prevented them from supporting its introduction. Government should provide adequate funding to upgrade labour rooms in a way that provides privacy to the delivering women and frame guidelines and train Birth Companions to perform their role appropriately.

## INTRODUCTION

Attempts to improve maternal and neonatal outcomes over the past two decades have emphasised conduct of deliveries in health care facilities, which have risen 54% globally from 50% (2000) to 77% (2020)^1^, while in India institutional births have more than doubled from 39% (2005) to 89% (2015).^2^ Maternal mortality ratio (MMR), in turn, has fallen in India by 55%, from 254 per 1,00,000 live births in 2004-06 to 113 per 1,00,000 live births in 2014-16. ^3^

Despite such obvious improvement in health indices, client satisfaction surveys of women delivering in public health institutions reveal an unsatisfactory scenario in several parts of the world, including in India where the satisfaction level is a meager 68%. ^4 5 6^ Many women undergoing institutional delivery in public health setups face undignified and disrespectful treatment from health care providers in the form of physical, verbal and emotional abuse. ^7 8 9^ Fear of being alone in unfamiliar surroundings, in contrast to ‘safe and reassuring environment’ at home, compels some women to deliver at home. ^10^ Presence of a Birth Companion (BC) with a woman in labour is known to improve beneficiary satisfaction and positively influence birth outcome in the form of shorter labour, better pain control and reduced need for medical interventions. ^11^ Respect for human rights including the right to self determination and privacy, which is embodied in the idea of Respectful Maternity Care (RMC), includes a companion of choice throughout labour.^12^ Based on results of a Cochrane systematic review of 22 trials, WHO, in 2015, recommended presence of BC during labour and childbirth. ^13^

A BC is any person chosen by a woman to provide her with continuous physical, emotional and psychological support during labour and childbirth.

Sufficient evidence exists that presence of a person of choice with the woman during labour and childbirth has a positive influence on overall birthing experience. A randomized controlled clinical trial in a regional teaching hospital in Thailand was conducted to evaluate the efficacy of a female BC.. ^14^ It found that women in experimental group with a female BC had a significantly shorter duration of active labour and were more satisfied with their childbirth experiences. Evidence states that disrespectful or abusive care during childbirth in facilities negatively influenced maternal and newborn outcomes ^15^ and thus there is emphasis on allowing a person of choice during childbirth. A WHO report^16^ states that women who are allowed a companion during childbirth report less fear and distress, which acts as a buffer against adverse aspects of medical intervention. Presence of a BC has clinically meaningful benefits in terms of shorter labour, increased rate of spontaneous vaginal birth, decreased use of intra-partum analgesia, decreased need for instrumental deliveries & cesarean section and increased satisfaction during the birthing experience; newborns of these women had a higher 5 minute Apgar scores. The above evidence was based on a Cochrane systemic review by Bohren et al who analyzed data pooled from 26 trials from 17 countries involving 15858 women. ^17^ Based on this evidence, WHO has recommended^16^ continuous companionship during labour and birth to improve women’s satisfaction with health care services.

Reviews have identified concern among the health care providers and also health system factors as barriers to BC. ^18 19^ A study from Sri Lanka has pointed to the need to improve awareness among the practitioners of the benefits of BCs. ^20^

A study in Tanzania concluded that health providers are the gatekeepers of companionship, ^21^ and facilities where providers experience staff shortages and high workload may be particularly responsive to programmatic interventions that aim to increase staff acceptance of Birth Companionship. A pilot project in Tanzania in nine Government health facilities and six comparison sites found that the introduction of birth companionship in participating facilities was feasible and well accepted by health providers, Government officials and most importantly, women who delivered at those facilities. ^22^

Many countries are including BC in their maternal health guidelines. Some countries, for example China, ^18^ Shri Lanka, Brazil ^20^ and Kenya^21^ have begun to allow a BC in their public health setups to certain extent. A national law in Brazil in 2005 affirmed the rights of all women to have a companion of choice during labour, delivery and postpartum period.^23^ National Guidelines for Quality Obstetrics and Perinatal Care in Kenya recommend a BC chosen by woman during first and second stage of labour. ^24^ Sri Lanka formulated a policy in 2011 to allow Birth Companions. ^20^

In India, Christian Medical College, Vellore in the state of Tamil Nadu started a birth companion programme in 2002. An order was issued by the State Government in July 2004 to scale up this programme for all Government hospitals in the state. Subsequently, Tamil Nadu witnessed a reduction in the maternal mortality ratio (MMR) from 380 in 1993 to 90 in 2007. ^25^ In India, although BCs are being allowed in some private hospitals, there are very few Government hospitals where this concept is being put to practice. Tamil Nadu was the first state to initiate BC scheme in all public hospitals. ^26^ The Ministry of Health & Family Welfare decided in February 2016 to allow BC to reduce maternal and infant mortality^27^ and launched Labour Room Quality Improvement Initiative (LaQshya)^28^ in March 2018 to implement this decision. The Ministry of Health and Family Welfare in India has also introduced the concept of BC under LaQshya programme - an intervention to improve maternal and neonatal outcomes^28^and to enhance satisfaction of beneficiaries in health facility and provide Respectful Maternity Care (RMC). Since approximately 46% maternal deaths, over 40% stillbirths and 40% newborn deaths take place on the day of the delivery itself, LaQshya aims to reduce preventable deaths where birth take place in labour room or maternity operation theater. It aims to provide an effective system to enhance satisfaction of beneficiaries and provide RMC to all pregnant women attending the public health facility. ^28^ Despite an advisory from the Health Ministry in February, 2016, ^27^ BC were not widely allowed in Government hospitals beyond Kerala, ^29^ Punjab^30^ and UP. ^31^

The advent of COVID-19 pandemic in March 2020 disrupted all aspects of daily life globally, including health care. Maternal and child health services suffered a setback.^32^ The WHO universally recognized that all pregnant women including those with confirmed or suspected COVID-19 infection have the right to high quality care, including the presence of companion of choice during delivery. ^33^ The role of birth companion has become all the more important than ever before as he / she can be utilized to minimize unnecessary patient-clinician interface and optimize manpower in this critical time.^34^ The Government of India has also acknowledged that the COVID-19 has had maximum impact on women, children and adolescents. It has reiterated women’s empowerment, so that they are able to make informed decisions around safe motherhood practices, including choice of birth companion. ^35^

Pandemic related stressors, worries and social distancing have affected the mental health of pregnant women, with a high percentage of them having symptoms of depression and anxiety.^36^ Therefore role played by a birth companion under these circumstances is of a higher magnitude than ever before.

Despite these recommendations, guidelines and laws, implementation of the concept of BC has not been optimal. Studies have found the implementation of BC in Sri Lanka as unsatisfactory. ^20^ A study on frequency of women having BC in Brazil found that approximately one quarter of women had no companion at all, less than one in five had continuous companionship, and 55.2% had partial companionship. ^23^ Even in the state of Tamil Nadu in India, where a Government order was passed to permit a BC in all public health hospitals, ^25^ many secondary and tertiary public hospitals and private hospitals do not allow a BC, while most Primary Health Centers (PHCs) do, making the women prefer PHCs for delivering their baby. ^37^ LaQshya program was launched in March, 2018 with an aim to achieve tangible results within 18 months that will benefit every pregnant woman and newborn delivering in public health institutions; ^38^ yet only 9% (262 of 2805 identified facilities) labour rooms had been nationally certified by November, 2020.^39^

Present literature reveals that the reasons for low implementation of BC concept are multi-factorial. A cross sectional study among practitioners in Sri Lanka identified lack of physical space in the labour room and the volume of work for non-adoption of BC. ^20^ According to a study conducted in Zambia, all the health staff stated that hospital policy was the principal reason for prohibiting companionship during labour. ^40^ They also had the apprehension that social support persons would interfere with their work by giving traditional medicines to women in labour.

A qualitative research was carried out in public teaching hospitals in three Arab countries. ^41^ Midwives and nurses pointed out structural factors while obstetricians pointed out social norms as barriers to implementation of the BC concept. To find women’s and provider’s perceptions on the issue of BC, a mixed-method study was done in Kenya. ^42^ It identified embarrassment, fear of gossip, abuse, privacy concerns, distrust of companion and ward set-up on women’s behalf as barriers. Providers who allowed BC stated that the companion could be useful to help providers with some chores, purchase supplies, hold flashlights and help in cleaning up.

We looked for studies which gave an insight into solutions to overcome barriers to implementation of the concept of BC. Researchers from Brazil stated that change in institutional culture and rules, such as having a clear policy, and changes in facilities such as having chairs for all companions were associated with implementation of BC. ^23^ Khasholian and Portela, after a review of 41 publications^19^ concluded that understanding providers’ attitude and sensitizing them was necessary before introducing companion of choice at birth. They recommended a committed management team which could change hospital policies.

The Guidance Development Group (GDG) members of WHO who developed the recommendations on the concept of BC suggested modification of physical space in facility, sensitization and training of health care workers to increase acceptance and orientation of the companions as measures to improve implementation. ^13^

We did a thorough literature review on studies from India on awareness about the concept of BC among health care providers, perceived barriers to its implementation and suggested solutions to overcome them. Except for a WHO report, ^26^ some news items, ^29 30 31^ and a couple of research papers^25 37^ which touched upon the topic of BC in general, we did not find, to the best of our knowledge, any scientific study relating to our study objectives.

During our literature search, we found lacunae in the existing knowledge on probable reasons for non-adoption of BC in public health facilities in India. Is the non adoption because of a lack of awareness among health care providers or because they face constraints in implementing it? Since this is a novel initiative, there is a possibility that health care providers in the field of obstetrics are not aware or only partly aware of the concept of BC. Health care providers are uniquely placed to pinpoint qualities most desirable in a BC, barriers to its implementation and ways to overcome the identified barriers. We decided to carry out this study with the objective of knowing the level of awareness about the concept of BC among health care providers in a tertiary level teaching institution in the capital city of India; whether the providers approved of allowing a companion at birth, barriers to implementation of this concept and ways to overcome them.

The aim of the study was to assess the level of awareness among the health care providers in a tertiary care hospital in North India of the concept of BC in labour and delivery, as also perceived barriers and suggestions for its implementation.

## METHODS

A facility based, cross-sectional quantitative study was conducted in the Department of Obstetrics & Gynecology, Lok Nayak Hospital, Maulana Azad Medical College (MAMC), New Delhi, India. Data was collected in the month of June and July 2019 after obtaining approval of the institutional ethics committee.

### Study Population and Sample Size

We interviewed all the health care providers involved in providing institutional care during labour and delivery, namely doctors (consultants, senior residents hereinafter called residents and Post-Graduate students) and nurses (Senior and Staff). There was no other inclusion criteria. Those who were unwilling to participate or did not give their consent were excluded. Universal total population sampling ^43^ was done, under which all individuals belonging to the study population were approached for participation. This was done to eliminate any bias or sampling error. Of 115 doctors, 96 were interviewed (response rate of 83%), while 55 of 105 nurses were interviewed (response rates of 52%). With a sample of 151, our overall response rate was 69 %. Two third of the respondents (64%) were doctors (10 consultants, 47 Post Graduates, 39 residents), with half being Post Graduates forming the largest group (31%), rest being nurses (15 senior nurses, and 40 staff nurses). Being an Obstetrics department, most (95%) of the respondents were females. There were no male nurses in the Department.

### Ethical Clearance

Institutional Ethics Committee of Maulana Azad Medical College accorded clearance to the conduct of the study (order No. F.17/IEC/MAMC/19/No. 125 Dated 27.5.2019). The option to opt out of the study was kept open without any condition. Complete confidentiality regarding participant’s information was maintained.

### Study Instrument

We framed the study questionnaire based on literature review on the perceived barriers to and possible solutions to introduction of BC during delivery. A structured, written questionnaire in English was pre-tested on 10 subjects, following which it was suitably modified and finalized (Annexure-3). Study Participant Information Sheet (Annexure-1) and Consent form (Annexure-2) were also prepared. The respondents were informed of the objectives and procedure of the study, informed consent obtained from each willing participant before administering the questionnaire. First three questions captured the socio-demographic characteristics and position of participants in the department. Questions 4-6 related to awareness of the concept of BC. Those not aware were briefly told about the concept so that they could answer the subsequent questions numbered 7-15.

### Data Analysis

The information collected was entered in a Google form, and results downloaded as a Comma Separated Values (csv) file. Data was checked for consistency and completeness; data entry errors were spotted and corrected. Cleaned up data was analyzed in Stata 8.0. Labels were assigned to variables. Ordinal variables were coded in a consistent, hierarchical manner with the lowest score of 1 denoting full awareness, highly beneficial or strong agreement.

The primary outcomes of this research were awareness of, opinion on the qualification of and benefits of, barriers to, suggestions and applicability of the concept of BC. Any systematic differences of outcome variables by demographic variables or position in the department were ascertained.

The study data contained responses of participants to various questions – either as binary, or on an ordinal scale of an order of three or five. Since ordinal variables and differences between them are neither uniform nor can be assumed to be normally distributed, standard measurements methods like means, standard deviations and t-test are not suitable^44^. Instead, we used non-parametric tests^45 46^ like Pearson Chi Square statistic, Fisher’s exact test where cell values were less than five, Kendall’s tau-b, Kruskal’s gamma statistic and Kruskal-Wallis test that do not rely on the assumption of normal distribution. For ordinal data, median and percentages were employed. Quantile regression was used for a similar reason. P-value of less than 0.05 was considered significant.

## RESULTS

Of the 151 participants, 143 (95%) were female. Age of the participants ranged from 21 to 55 years, with a mean age of 31 years. (Table 1)

**Table 1:** Demographic Characters of Respondents

| Position | Numbers (%) | Mean age | Female | Male |
| --- | --- | --- | --- | --- |
| Consultant | 10 (6.62 ) | 47 | 9 | 1 |
| PG | 47 (31.13 ) | 26 | 42 | 5 |
| Resident | 39 (25.83 ) | 29 | 37 | 2 |
| Sr. Nurse | 15 (9.93 ) | 43 | 15 | 0 |
| Staff Nurse | 40 (26.49 ) | 32 | 40 | 0 |
| Total | 151 (100) | 31 | 143 | 8 |

Awareness of the concept of BC (Q. 4) and WHO recommendation to that effect (Q. 5) was very high, with 53% (n=80) and 54% (n=82) of respondents respectively being fully aware, 40% (n=61) and 29% (n=44) respectively being somewhat aware (Table-2). Median awareness for both questions was 1 (Table-3), denoting full awareness on a three point scale with 3 being not aware. Sub-group analysis revealed highest level of awareness among consultants (Table-3, mean and median awareness was 1 and close to 1), and a high level of awareness among staff nurses (median awareness 1). We used non-parametric tests on the ordinal responses to the two sets of questions on awareness to find their association. (Table-2)

**Table-2:** Awareness of WHO Recommendation Regarding Birth Companion

| **Q4. Are you aware of the concept of birth Companion ?** | **Q5. Are you aware that the WHO recommends every woman to be accompanied by a Birth companion ?** |  |  |  |
| --- | --- | --- | --- | --- |
|  | Fully Aware | Somewhat aware | Not aware | Total |
| Fully Aware | 57 | 16 | 7 | 80 |
|  | 71.25 | 20 | 8.75 | 100 |
|  | 69.51 | 36.36 | 28 | 52.98 |
| Somewhat Aware | 25 | 27 | 9 | 61 |
|  | 40.98 | 44.26 | 14.75 | 100 |
|  | 30.49 | 61.36 | 36 | 40.4 |
| Not Aware | 0 | 1 | 9 | 10 |
|  | 0 | 10 | 90 | 100 |
|  | 0 | 2.27 | 36 | 6.62 |
| Total | 82 | 44 | 25 | 151 |
|  | 54.3 | 29.14 | 16.56 | 100 |
|  | 100 | 100 | 100 | 100 |

Pearson chi2(4) = 55.6358 Pr = 0.000
Cramér's V = 0.4292
gamma = 0.6271 ASE = 0.091
Kendall's tau-b = 0.4023 ASE = 0.072
Fisher's exact = 0.000

1. Pearson Chi Square statistic (χ2) with 4 degrees of freedom is 55.6 with a p-value of 0.00, denoting that the observed responses in rows and columns to the two questions are associated, dependent and significantly related, rejecting the null hypotheses of independence.
2. Since some of the cells had values below five, violating the assumption of Chi-Square Test, we used Fisher’s exact test, which also showed dependence of the response to the two questions (p=0.00).
3. Since both dimensions of the table can be ordered, Kendall’s tau-b coefficient can be used to test the strength of association of cross tabulations, which has an Alpha’s Standard Error (ASE) of 0.072 denoting dependence of two sets of responses.
4. Kruskal’s gamma statistic at 0.62 and ASE of 0.091 also show a material level of association.

On the question of awareness of Government’s guideline on the presence of BC (Q.6), awareness levels were slightly lower with 39% (n=59) being fully aware, while 29% (n=44) were somewhat aware (Table-4) and an overall median score of 2 (Table-3). Consultants were still highly aware of this stipulation (Median 1, mean 1.3), while senior nurses were least aware of it (Median 3, mean 2.2).

**Table-3:**
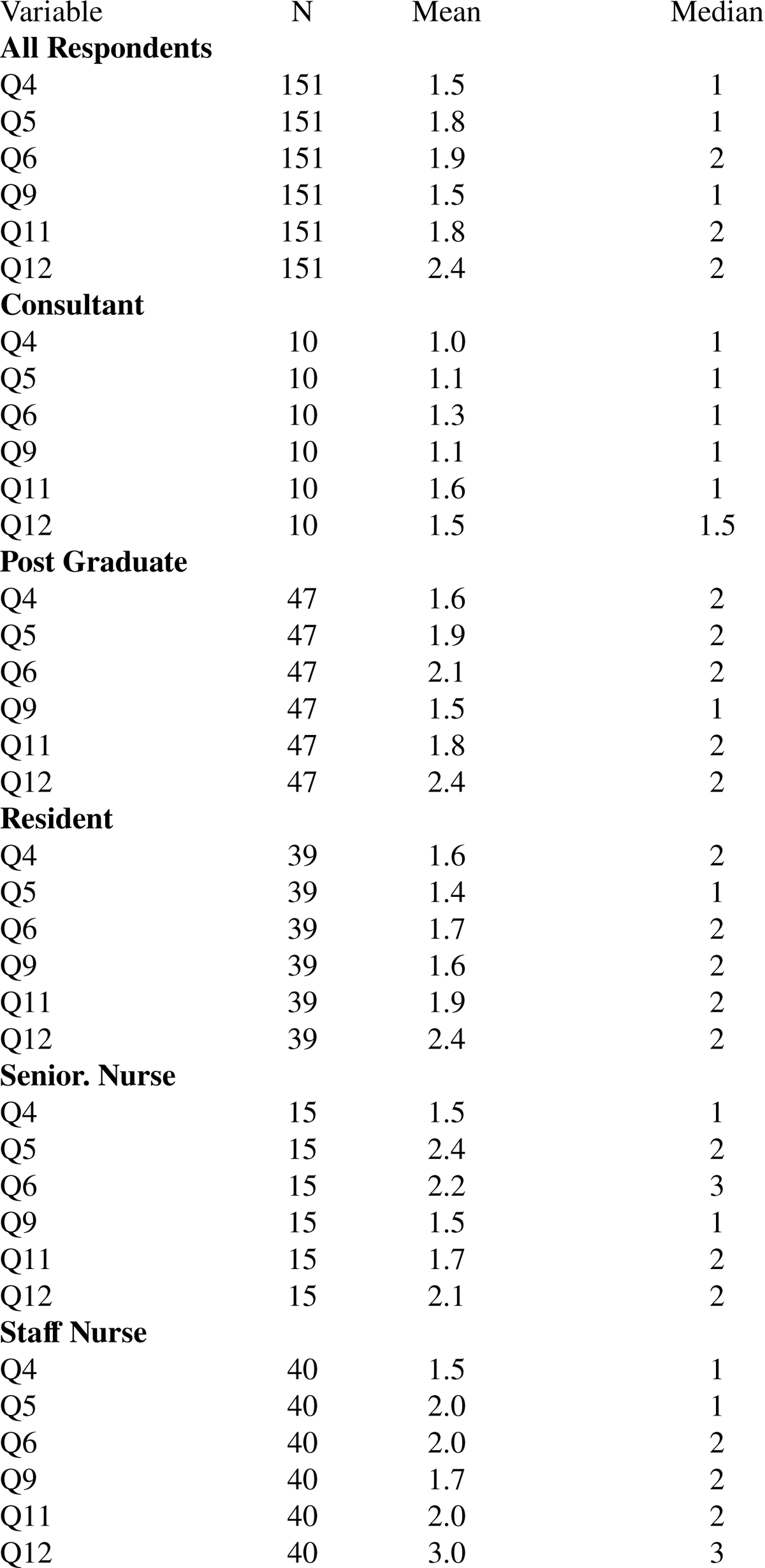
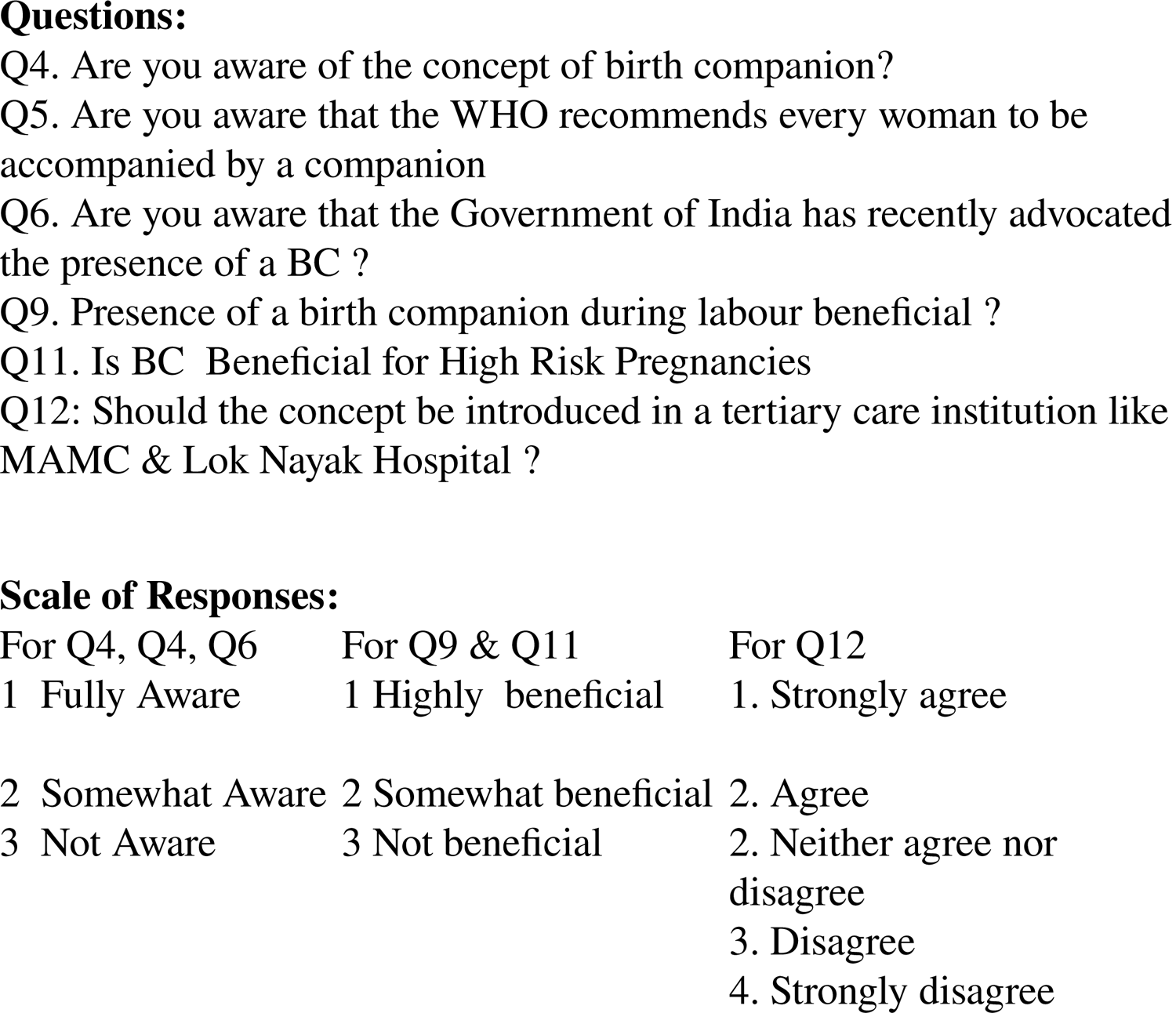
Awareness of BC concept, perception of its benefit with Sub-Group responses

**Table-4:** Awareness of Birth Companion vs. that of Government Guidelines

| Q4. Are you aware of the concept of birth Companion ? | 6. Are you aware that the Government of India has recently advocated the presence of BC in all government hospitals ? |  |  |  |
| --- | --- | --- | --- | --- |
|  | Fully Aware | Somewhat aware | Not aware | Total |
| Fully Aware | 45 | 20 | 15 | 80 |
|  | 56.25 | 25 | 18.75 | 100 |
|  | 76.27 | 45.45 | 31.25 | 52.98 |
| Somewhat Aware | 13 | 23 | 25 | 61 |
|  | 21.31 | 37.7 | 40.98 | 100 |
|  | 22.03 | 52.27 | 52.08 | 40.4 |
| Not aware | 1 | 1 | 8 | 10 |
|  | 10 | 10 | 80 | 100 |
|  | 1.69 | 2.27 | 16.67 | 6.62 |
| Total | 59 | 44 | 48 | 151 |
|  | 39.07 | 29.14 | 31.79 | 100 |
|  | 100 | 100 | 100 | 100 |

Of those who were fully aware about the concept of BC, 71% were also fully aware of the WHO recommendation (Table-2) and 56% were fully aware that Government of India advocates presence of BC (Table-4). The association to the responses of awareness of the concept of BC and Government’s advisory was significant with a Pearson Chi Square statistic (χ2) with 4 degrees of freedom of 29.6 (p-value: 0.00), Fisher’s exact test p-value of 0.00, Kendall’s tau-b of 0.37 (ASE: 0.067) and Kruskal’s gamma statistic at 0.37 and ASE of 0.067 (Table-4).

Similarly, the responses to the question on WHO’s recommendation and Government’s advisory were significantly related with a Pearson Chi Square statistic (χ2) with 4 degrees of freedom of 64.5 (p-value: 0.00), Fisher’s exact test p-value of 0.00, Kendall’s tau-b of 0.56 (ASE: 0.054) and Kruskal’s gamma statistic at 0.78 and ASE of 0.057 (Table-5).

**Table-5:** Awareness of WHO recommendation vs. Government Guidelines

| Q5. Are you aware that the WHO recommends every woman to be accompanied by a Birth companion ? | Q6. Are you aware that the Government of India has recently advocated the presence of BC in all government hospitals ? |  |  |  |
| --- | --- | --- | --- | --- |
|  | Fully Aware | Somewhat aware | Not aware | Total |
| Fully Aware | 51 | 22 | 9 | 82 |
|  | 62.2 | 26.83 | 10.98 | 100 |
|  | 86.44 | 50 | 18.75 | 54.3 |
| Somewhat Aware | 7 | 19 | 18 | 44 |
|  | 15.91 | 43.18 | 40.91 | 100 |
|  | 11.86 | 43.18 | 37.5 | 29.14 |
| Not Aware | 1 | 3 | 21 | 25 |
|  | 4 | 12 | 84 | 100 |
|  | 1.69 | 6.82 | 43.75 | 16.56 |
| Total | 59 | 44 | 48 | 151 |
|  | 39.07 | 29.14 | 31.79 | 100 |
|  | 100 | 100 | 100 | 100 |

Respondents were asked to rank the preferred BC from a list (Q.7). Mother was the BC of choice (n=106, 70%) closely followed by husband (n=104, 69%, Table 6). The pre-requisite for being a BC were identified as wearing clean clothes (95%, n= 144), not suffering from any communicable disease (91%, n=138), staying with the woman throughout the process of labour (74%, n=111). A minority of respondents opted for the BC having gone through the process of labour (42% n=64), being a female relative (40%, n=61). Only 15% (n=22) responded opined that BC should attend to other women in the labour room, while only 7% respondents (n=10) thought that BC would interfere with the work of hospital staff (Table 7).

**Table 6:** Responses to Who can be a Birth Companion.

| Variable | N | Mean | % |
| --- | --- | --- | --- |
| Mother | 151 | 106 | 70% |
| Husband | 151 | 104 | 69% |
| Sister | 151 | 69 | 46% |
| Nurse | 151 | 65 | 43% |
| Mother-in-law | 151 | 52 | 34% |
| Friend | 151 | 46 | 30% |
| Doctor | 151 | 43 | 28% |
| Paid Daula | 151 | 15 | 10% |
| Children | 151 | 1 | 1% |

Note: Since more than one choice was allowed, sum exceeds 151

**Table 7:** Pre-requisite for a Birth Companion

| Pre-requisites for a birth companion ? | N | Agree | % |
| --- | --- | --- | --- |
| 8.4 She should wear clean clothes | 151 | 144 | 95% |
| 8.3 Should not suffer from any communicable disease | 151 | 138 | 91% |
| 8.5 She should stay with the pregnant woman through the process of labour | 151 | 111 | 74% |
| 8.2 She should have gone through process of labour ? | 151 | 64 | 42% |
| 8.1 Should be female relative. | 151 | 61 | 40% |
| 8.7 She should attend to other women in the labour room | 151 | 22 | 15% |
| 8.6 She should not interfere with the work of hospital staff | 151 | 10 | 7% |

On the perceived benefits of a BC during labour (Q.9), most respondents (n=51%, n=77) viewed it to be highly beneficial, many (n=44%, n=67) felt it was somewhat beneficial (Table-10) with a overall median score of 1 (Table-3). Most consultants, Post-Graduates and Senior Nurses opined it to be highly beneficial with a median score of 1. Residents and Staff Nurses were muted in their response and believed BC would be somewhat beneficial (median score: 2, Table-3).

Respondents were then asked (Q.10) of the likely benefits of a BC (Table-8). Almost everyone (99%, n=149) opined that BC would provide emotional support and boost the woman’s confidence. More than 90% participants opined that the BC would provide comfort measures like soothing touch, massage, increase satisfaction, spiritual support, early initiation of breastfeeding, help the woman to advocate her wishes to others, and reduce postpartum depression. Majority thought that it would lead to humanization of labour, reduced need for analgesia, increased spontaneous vaginal births, reduced incidence of unnecessary cesarean sections, reduced need for instrumental delivery, reduced workload for hospital staff and in delivering in birth position of choice. Half the respondents thought that it would lead to a higher Apgar score. Some respondents expected BC to lead to a shorter duration of labour (38%), or reduced intra-partum bleeding (36%) or increased use of partograph (26%, Table-8).

**Table-8:** Perceived benefits of a Birth Companion

| Variable | N | Agree | % |
| --- | --- | --- | --- |
| 10.10 Emotional support | 151 | 149 | 99% |
| 10.14 Boosting woman's confidence | 151 | 148 | 98% |
| 10.11 Comfort measures – soothing touch, message | 151 | 144 | 95% |
| 10.16 Increased satisfaction by women | 151 | 142 | 94% |
| 10.15 Spiritual support | 151 | 141 | 93% |
| 10.18 Early initiation of breastfeeding | 151 | 141 | 93% |
| 10.12 Helping the woman to advocate her wish to others | 151 | 140 | 93% |
| 10.20 Reduced post-partum depression | 151 | 137 | 91% |
| 10.6 Humanization of labour | 151 | 126 | 83% |
| 10.2 Reduced need for analgesia. | 151 | 106 | 70% |
| 10.3 Increased spontaneous vaginal births. | 151 | 104 | 69% |
| 10.5 Reduced incidence of unnecessary cessarian sections | 150 | 98 | 65% |
| 10.4 Reduced need for instrumental delivery | 151 | 92 | 61% |
| 10.7 Reduced workload for hospital staff. | 151 | 86 | 57% |
| 10.13 Delivering in birth position of choice | 151 | 82 | 54% |
| 10.19 Higher newborn Apgar score | 151 | 75 | 50% |
| 10.8 Avoid frequent vaginal examination | 151 | 60 | 40% |
| Q10_1 benefits-shorter duration of labour | 151 | 58 | 38% |
| 10.9 Reduced intrapartum bleeding | 151 | 54 | 36% |
| 10.17 Increased use of partograph | 151 | 39 | 26% |

Most respondents thought that BC would be somewhat beneficial in dealing with high risk pregnancies (median response: 2, Q.11). Most consultants, however, viewed it be highly beneficial with a median score of 1 (Table-3).

Seeking readiness for practice, respondents were asked if the concept of BC should be introduced in a tertiary care institution like theirs, MAMC & Lok Nayak Hospital (Q.12, Graph-1, Graph-2 Table-9). Respondents strongly agreeing to this suggestion was only 19% (n=29) with 40% (n=60) agreeing. The median response was “Agree” (mean: 2.4, median: 2 on a scale of 1 to 5, Table-3, 10, Graph-1). This response was milder than to previous question on expected benefits of a BC (Q.9) where the overall perception was of being “highly beneficial” (Table-3). 75% of respondents who reported the BC to be highly beneficial agreed to the suggestion of it being implemented in their hospital (Table-9). A smaller percentage of respondents (45%) who observed BC to be somewhat beneficial agreed to its introduction in their hospital (Table-9). The responses to the question on expected benefits of BC and introducing in their hospital were significantly related with a Pearson Chi Square statistic (χ2) with 8 degrees of freedom of 38.9 (p-value: 0.00), Fisher’s exact test p-value of 0.00, Kendall’s tau-b of 0.37 (ASE: 0.064) and Kruskal’s gamma statistic at 0.56 and ASE of 0.08 (Table-9).

**Table-9:**
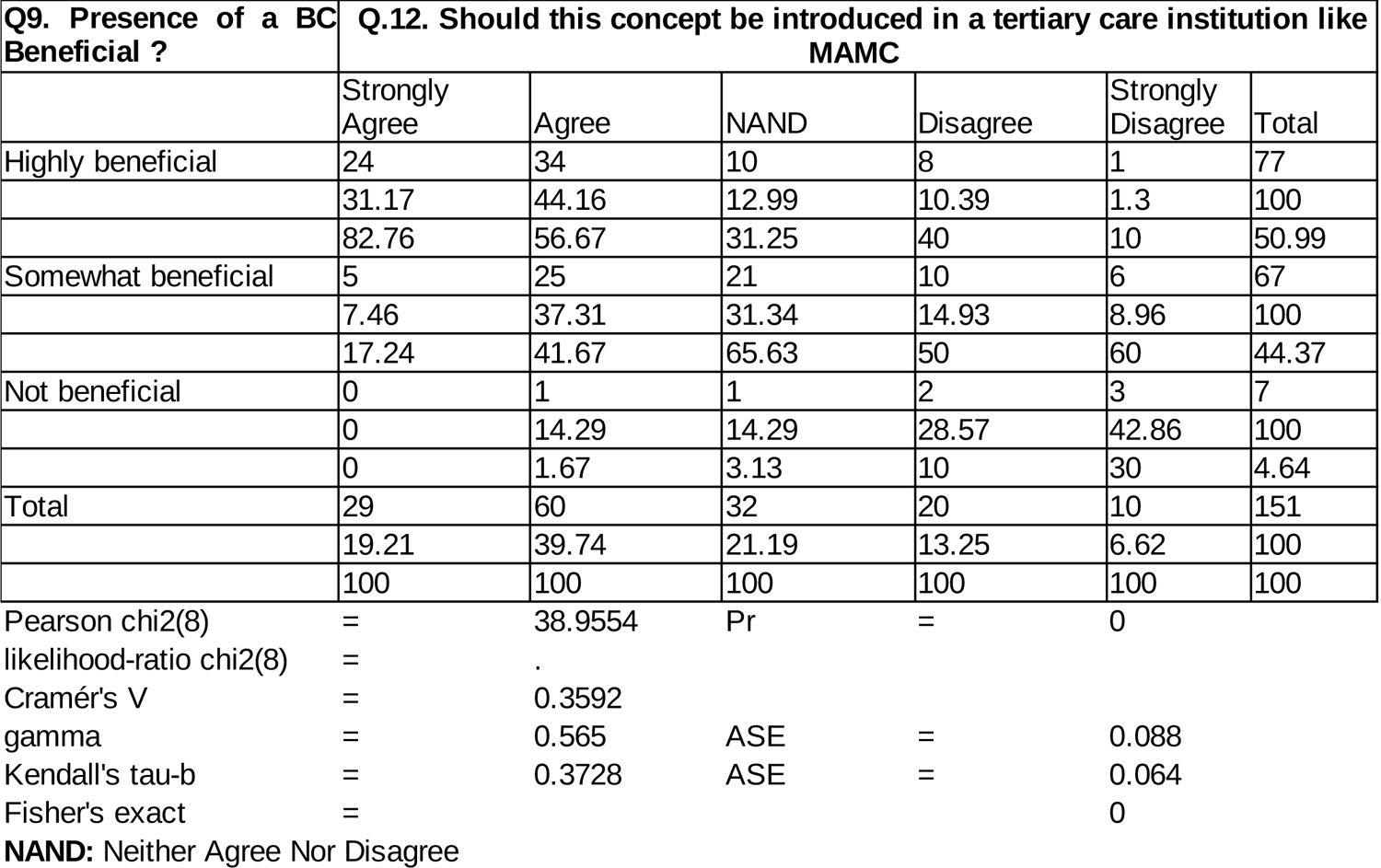
Responses to Benefits of BC vs. Introducing it in their Hospital

Among groups of respondents, consultants were the most supportive (median and mean: 1.5 on a five point scale where 1 is strongly agree, 5 strongly disagree, Table-3, Graph-2) while staff nurses were the least approving (mean: 3, median: 3) of introducing BC in their hospital. Kruskal–Wallis one-way analysis of variance to test the null hypothesis that the medians of all groups are equal (Table-11) is rejected with Chi-Square value of 16.8, p-0.001. Using quantile regression which estimates the median of the dependent variable, conditional on the values of the independent variable, we find that as we move lower in the health care worker hierarchy, the support for the introduction of BC in their hospital falls. (coefficient: 0.33, t: 5.59, p: 0.00, Table-12). Thus, response to the question on introduction of BC in their hospital was dependent on position, with least support among the staff nurses.

**Table-10:** Opinions on applicability of, barriers to, suggested corrective measures for introduction of Birth Companion in hospitals

| Variable | N | Mean | Median |
| --- | --- | --- | --- |
| 12. Should this concept be introduced in a tertiary care institution like MAMC ? | 151 | 2.5 | 2 |
| Q13_1_1 Barriers At the Institution Level -Hospital Policy | 150 | 2.3 | 2 |
| Q13_1_2 Over-crowding in Labour Room | 151 | 1.4 | 1 |
| 13.1.3: Lack of privacy for the woman (no dividers or curtains) | 151 | 1.9 | 2 |
| 13.1.4 Privacy concerns for other women, especially in presence of male companion | 151 | 1.5 | 1 |
| 13.1.5 Risk of infection transmission | 151 | 1.8 | 2 |
| 13.1.6 Asepsis in OT will be compromised | 149 | 2 | 2 |
| 13.1.7 Women would not cooperate with hospital staff in presence of birth companion | 150 | 2.5 | 2 |
| 13.1.8 Use of traditional medicines by birth companion | 150 | 2.5 | 2 |
| 13.1.9 Birth companion may interfere with clinical decision making | 150 | 2.3 | 2 |
| 13.1.10 A lounge / waiting area will be needed for the birth companion to take | 150 | 2 | 2 |
| 13.1.11 The birth companion may not be continuously present during labour | 150 | 2.3 | 2 |
| 13.1.12 Birth companion will not be able to see the sight of blood | 149 | 2.2 | 2 |
| 13.1.13 Woman's health information should be kept confidential | 150 | 2.1 | 2 |
| 13.1.14 Presence of birth companion is unprofessional | 150 | 3.2 | 3 |
| 13.2.1 For the patient: Embarrassment of delivering before someone else | 150 | 2.6 | 2 |
| 13.2.2 Not socially acceptable (stigma / customs) | 150 | 2.9 | 3 |
| 13.2.3 Birth companion may gossip in the community | 150 | 2.5 | 2 |
| 13.2.4 Cost of transportation of birth companion | 150 | 2.8 | 3 |
| 13.2.5 Economic loss if birth companion is an earning member and has to take br | 150 | 2.5 | 2 |
| 13.2.6 Hiring a duala is expensive | 151 | 2.3 | 2 |
| 13.2.7 No trustworthy person to accompany her as a birth companion | 150 | 2.6 | 2 |
| 14.1 How can these barriers be overcome ? - Increased awareness among the hospital staff | 151 | 1.8 | 2 |
| 14.2 creating physical partition to ensure privacy | 151 | 1.6 | 2 |
| 14.3 providing incentives to women delivering in the presence of birth companion | 151 | 2.8 | 3 |
| 14.4 prior training of birth companion on their role | 151 | 2 | 2 |
| 15.1 Provide funding to hospitals to upgrade labour rooms | 151 | 1.5 | 1 |
| 15.2 incentivize hospitals that allow birth companion | 151 | 2.2 | 2 |
| 15.3 formulate guidelines for instructing birth companion | 151 | 1.7 | 2 |

**Table-11:** Kruskal-Wallis test on the Response by Position to Question on Whether BC concept be introduced in their Hospital

| position | Obs | Rank Sum |
| --- | --- | --- |
| Consultant | 10 | 372.5 |
| PG | 47 | 3462.5 |
| Resident | 39 | 2920 |
| Sr. Nurse | 15 | 933.5 |
| Staff Nurse | 40 | 3787.5 |

chi-squared = 16.800 with 4 d.f.
probability = 0.0021
chi-squared with ties = 18.299 with 4 d.f.
probability = 0.0011

**Table-12.** Quantile Regression of Response on Introduction of BC in their Hospital by Position (Q.12)

|  |  |  |  |  |  |  |
| --- | --- | --- | --- | --- | --- | --- |
| Iteration 1: WLS sum of weighted deviations |  | = | 131.15488 |  |  |  |
| Iteration 1: sum of abs. weighted deviations |  | = | 132 |  |  |  |
| Iteration 2: sum of abs. weighted deviations |  | = | 126.66667 |  |  |  |
| Iteration 3: sum of abs. weighted deviations |  | = | 126.66667 |  |  |  |
| Median regression |  | Number of obs | = | 151 |  |  |
| Raw sum of deviations |  | 131 (about 2) |  |  |  |  |
| Min sum of deviations |  | 126.6667 | Pseudo R2 | = | 0.0331 |  |
| Q12 | Coef. | Std. Err. | t | P>t | [95% Conf. Interval] |  |
| Position | .3333333 | .0596357 | 5.59 | 0.000 | .2154925 | .4511742 |
| _cons | 1.333333 | .2001573 | 6.66 | 0.000 | .9378199 | 1.728847 |

**Graph-1:**
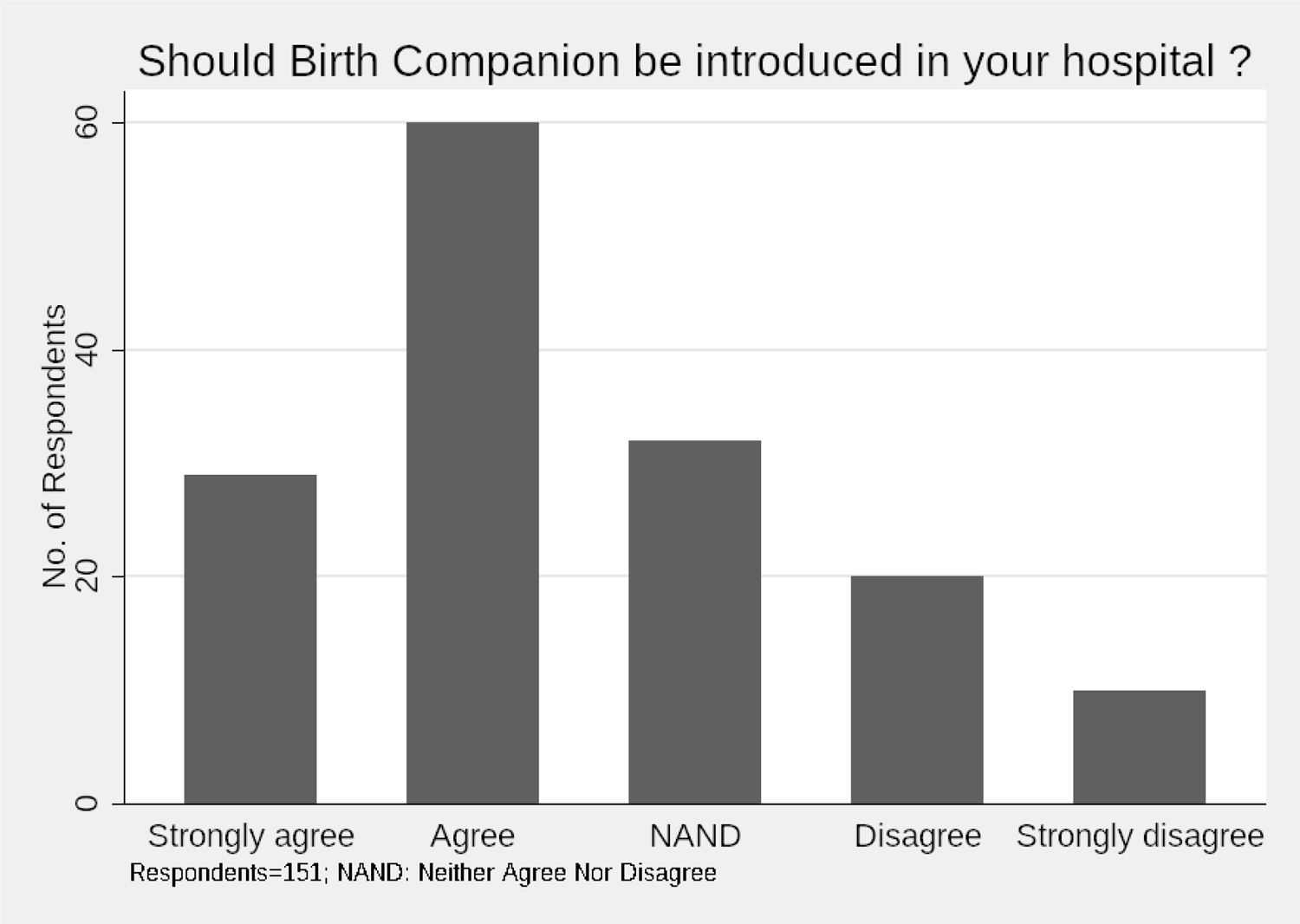
Response to Q.12 on introduction of Birth Companion

**Graph-2:**
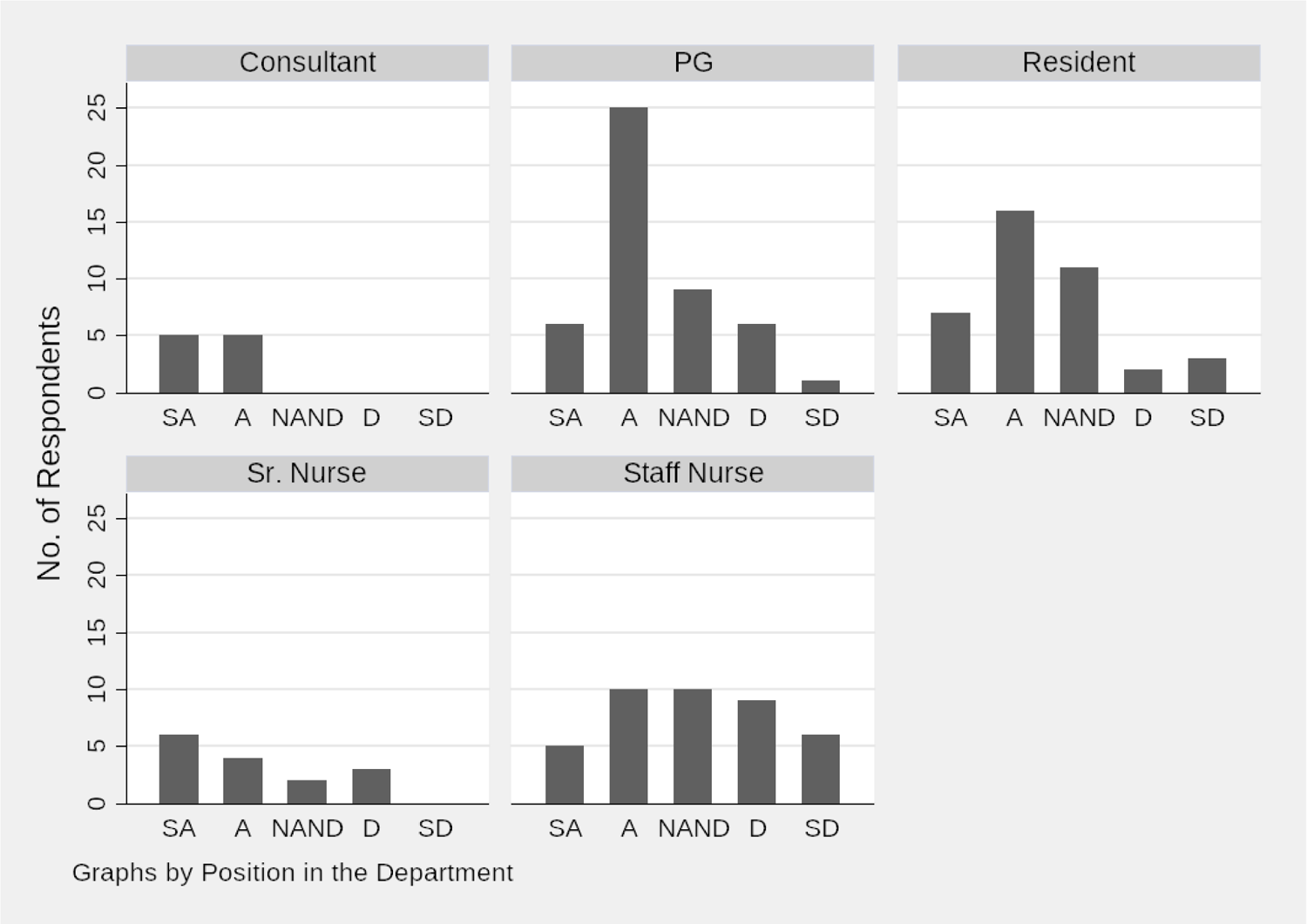
Histogram by Position of the Responses to Question 12 on Whether BC Should be Introduced in their Hospital

On possible barriers at institution level to introduction of a BC (Q. 13), there was a strong agreement on over-crowding in labour room and privacy concerns for other women, especially in presence of male companion (median score: 1, Table-10). Most respondents also ‘agreed’ with the suggestion that hospital policy could be a barrier, it posed a risk of infection transmission, asepsis in OT could be compromised, women may not cooperate with hospital staff in presence of BC, they may use traditional medicines, or may interfere with clinical decision making; that a lounge or waiting area will be needed for the BC, who may not be continuously present during labour or be able to see the sight of blood and that it may compromise the confidentiality of woman’s health information (median score: 2, Table-10). Most respondents were neutral (neither agreed nor disagreed) when asked if presence of BC was unprofessional (Table-10). On the patient side barriers agreed to included embarrassment for the patient of delivering in front of someone else, BC gossiping in the community, economic loss when BC is an earning member and expense of hiring a doula or lack of a trustworthy person (median score: 2, Table-10)

On the ways of overcoming the perceived barriers, the suggestion of providing funding to hospitals to upgrade labour rooms was strongly agreed to (median score 1, Table-10). Other options for overcoming the barriers agreed to were: increased awareness among hospital staff about benefits of BC, creating physical partition to ensure privacy, prior training of BC on their role, incentivising hospitals that allow BC and formulation of guidelines for instructing BC (median score: 2, Table-10). Most respondents were neutral (neither agree nor disagree) when asked if providing incentives to women who deliver in presence of a BC would help in overcoming barriers to implementation of the concept of BC (Table-10).

## DISCUSSION

Our respondents were representative of the team of doctors and nurses that manage labour and deliveries in the study institution. A wide range of respondents age (21 to 55 years) across health care provider categories underlines the diverse and rich experience that they bring to the study. A consensus or broad agreement among such a diverse group of care-givers signifies a widely held view and therefore carries high credibility, and generalizability.

Our study participants were largely aware of the concept of BC, and WHO recommendation that that every woman in labour and delivery should be accompanied by a companion of her choice. Two in every three (68%) of the interviewed health care providers were aware of Government of India stipulation on the presence of BC in all its hospitals. Awareness of WHO recommendation and Government of India stipulation among those who were already aware of the concept of BC was high (71%, 56% respectively), but provided scope for further improvement. Information sessions for health care providers should be organized to familiarize them on the guidelines of LaQshya and need for BC in the interests of quality of care, and dignity of the delivering women.

Even though the choice of a BC lies with the woman undergoing delivery, health care providers in our study identified mother and husband to be the most preferred BCs. These findings are similar to those of a study in Nigeria where husband was the BC of choice^47^ Policy and systems for allowing BC should accommodate this preference by allowing sufficient privacy for other women delivering in the same ward from the presence of a male attendants.

The pre-requisites identified for a BC include basics like hygiene, being disease-free and availability. While introducing the concept, screening of the BCs for communicable disease and a coaching on maintaining hygiene should be organized, as it will benefit the lady delivering and also neighbors. Being a woman, or having experienced labour were identified as pre-requisites for being a BC by over a third of respondents, showing willingness among the health care providers to accommodate the choice of the delivering woman.

High current levels of awareness of the concept of BC were matched by agreeing to its expected benefits (95% of respondents). Expected benefits particularly for high risk pregnancies were perceived to be marginally lower, but still a high proportion of respondents (85%). Consultants, who manage such cases, were largely in agreement of benefits in such cases. These findings are consistent with those from a study in Kenya, ^42^ where most providers recommended BC. Our findings are also aligned to those from a study from Sri Lanka where most of the respondent faculty members thought that presence of BC would provide moral support to women. ^16^

In our study 70% of respondent service providers felt that presence of BC would reduce the need for analgesia. This has been proven in a study by Marzieh et al in 2014^48^ which demonstrated that presence of doula during labour significantly reduced intensity of labour pain compared to the control group. Similarly, McGrath & Kennell found decreased need for epidural analgesia during labour among middle income women delivering with continuous labour support of a doula. ^49^

Most health care providers agreed to the suggestion that BC would provide emotional support, boost the woman’s confidence, provide physical, psychological and emotional comfort, and improve communication signifying an understanding of the mechanism by which BC can help the woman in labour. Almost 93% respondents opined that BC will lead to early initiation of breastfeeding, which has huge implications for child survival and post natal recovery. On the whole, health care providers perceived all round benefits of the presence of a BC.

Despite such high levels of awareness of the concept, usefulness and benefits of BCs, only 61% (88 of 144) respondents who perceived BC to be beneficial in Q.9 agreed to get it introduced in their hospital. Staff nurses were the least agreeable, with only 15 of 37 (40%) who agreed that BC could be beneficial in response to Q.9 agreed to its introduction in their hospital. The reasons for this steep difference between awareness and willingness to practice, and amongst health care providers points to practical difficulties in making the concept operational. These difficulties are captured in response to the questions on barriers and how these can be overcome.

Health care respondents across all categories strongly agreed that the key barrier to introducing BC in their hospital was overcrowding and privacy concerns for other women. Privacy concern for other women is similar to the response of health care providers given in the study from Kenya by Patience et al. ^42^ Inadequate space was also brought out as the most important barrier in an earlier study in Sri Lanka. ^20^

Other barriers agreed to by most heath care providers include the risk of infection transmission, compromising of asepsis in OT, hospital policy, need for a lounge or waiting area, apprehension of continued presence of BC or not being able to see the sight of blood, or compromising the confidentiality of woman’s health information. Presence of a BC was not perceived to be unprofessional. Lack of physical space leading to privacy issues as the key constraint in introducing BCs was supported by funding for hospitals emerging as the most preferred response to question on overcoming these barriers. Other suggestions included raising awareness of hospital staff and incentivising hospitals that allow BC. Our findings are consistent with WHO’s recommendation on the need for implementing a strategy for sensitization of health professionals, community and women towards acceptance of BC. ^13^

Apprehensions among health care providers that patients may not cooperate in the presence of BC, or BC may interfere in clinical decision making were marginal concerns, which can be overcome by formulating guidelines on conduct of BCs, and providing prior training to BCs. Cost of transporting BC was seen as non-issue as also the recommendation to provide incentives to women delivering in the presence of a BC.

Strengths of our study are – universal total sampling, reducing selection bias, and by interviewing all cadres of health care providers, we obtained a better understanding of the issue.

Our study had the following limitations – 1) Findings of our study are generalizable to tertiary care teaching institutions only, not to all public health facilities. 2) In the initial questions of the questionnaire, we tested awareness of the participants about the concept of BC. Those who were not aware were told about the concept in brief by the interviewer, while others were not briefed. This may have influenced answers to subsequent questions by the participants.

## Conclusion

In general, health care providers were largely aware about the concept of BC, WHO’s recommendation and Government of India (GoI) guideline on the presence of BC. Awareness on all these three areas, however, needs to be raised further among PG students, senior residents and staff nurses.

Most (95%) of service providers in this study found the BC to be highly beneficial or somewhat beneficial. Most participants identified mother and husband as the preferred BC of choice. Most respondents opined that BC would help in providing emotional support, increase satisfaction, help the pregnant woman to convey her wishes to others, initiate breast feeding early and reduce post partum depression. Birth Companions, therefore, can materially improve maternal and neonatal outcomes – a finding with with immense public health implications which is particularly relevant in the face of COVID-19, which has caused a setback to maternal and child health as well as lead to an acute shortage of health care staff. Barriers to introduction of BCs need to be identified and overcome.

Suggestions and Recommendations

Based on study findings, we offer the following suggestions.

1. Concept of BC should be part of teaching curriculum for PG and nursing students, which should be updated from time to time through Continuing Medical Education (CME) and Continuing Nursing Education (CNE).
2. All cadres of staff working in labour rooms should be briefed about LaQshya program under which the GoI advocates the presence of a BC.
3. Pregnant women should be informed during antenatal visits of benefits of a BC, and their right to carry a BC of choice at the time of delivery.
4. BC identified by the pregnant woman should be encouraged to accompany her for antenatal visits. The identified BCs should be screened and counseled so that they prepare themselves for this role.
5. Administrative heads of health facilities should should take concrete steps to facilitate presence of BCs. Available funds should be used to provide temporary partitions or curtains to create privacy for the women and their BC during delivery and labour.
6. India’s prescribed National Quality Assurance Standards for Government facilities already require (Standard B3) that the facility maintains privacy, confidentiality & dignity of patient. This needs to be implemented diligently by providing adequate funding to increase space, and provide privacy to women in labour and delivery. Hospitals that allow BCs should be incentivized.
7. Theory of Change^13^ that defines long-term goals and then maps backward to identify necessary preconditions can be used in hospitals to identify gaps, barriers and make a time-bound strategy to overcome them. Key stakeholders need to be engaged while drawing the action plan, and for its effective implementation. It is known that enhanced stakeholder involvement contributes to the quality of the services provided. ^50^

## Declarations

### Availability of data and materials

The data for this project is being shared as csv file.

### Author Contributions

TS conceived and designed the study, prepared the instrument, administered it and wrote the paper. YS and ST provided inputs on draft versions of the proposal, study protocol and drafts of the paper. RS helped in statistical analysis and wrote the conclusions. All authors read and approved the final manuscript.

## Data Availability

The data supporting the findings of this study are available within the article [and/or] its supplementary material.

## Acknowledgments

The authors will like to express sincere gratitude to the respondent heath care providers in sparing their time for responding to our questionnaire.

## Tables and Graphs

## Annexure-1

Maulana Azad Medical College and Lok Nayak Hospital, New Delhi —110 002 (Department of Obstetrics & Gynecology)

<u>STUDY PARTICIPANT INFORMATION SHEET</u>

You are being invited to participate in a research study.

Before you take part in this research study, the study must be explained to you and you must be given the chance to ask questions. Please read carefully the information provided here. if you agree to participate, please sign the informed consent form. You will be given a copy of this document to take home with you.

### STUDY lNFORMATlON

Protocol Title:

Awareness regarding, barriers to and suggestions for implementation of Birth Companion in labour and delivery: A cross sectional study among health care providers in a tertiary level teaching hospital in Delhi, India.

Principal Investigator(s):

……………… MBBS Student &

………………, MAMC.

PURPOSE OF THE RESEARCH STUDY

You are being invited to participate in a research study of Awareness, Barriers to and Suggestions for allowing Birth companion during labour and delivery. We hope to learn the reasons for non adoption of Birth Companion in tertiary level teaching institutions in India. You were selected as a possible subject in this study because you are a part of the team of health care providers in Department of Obstetrics & Gynaecology at a tertiary teaching hospital.

This study will recruit all the health care providers in Department of Obstetrics & Gynaecology at MAMC, which is a tertiary teaching hospital, for over a period of two months beginning June, 2019. About <u>137 respondents</u> will be involved in this study.

The study does not involve taking any sample of tissues, blood and/or body fluids. The study does not involve or provide access to any study medication/device.

STUDY PROCEDURES AND VISIT SCHEDULE

If you agree to take part in this study, you will be asked to answer a questionnaire. Your participation in the study will last about 10 minutes. You will not need to visit any place other than your work place at any time in the course of the study.

YOUR RESPONSIBILITIES IN THIS STUDY

If you agree to participate in this study, you should (Choose applicable points):

• Answer the questionnaire for the study.

WITHDRAWAL FROM STUDY

You are free to withdraw your consent and discontinue your participation at any time without any prejudice to you. if you decide to stop taking part in this study, you should tell the Principal Investigator. There are no adverse effects to you from possible withdrawl from this study.

The Principal Investigator of this study may stop your participation in the study at any time for one or more of the following reasons:

– Failure to follow the instructions of the Principal Investigator
– The study is cancelled.
– Other administrative reasons.

Unanticipated circumstances.

WHAT IS NOT STANDARD CARE OR EXPERIMENTAL IN THIS STUDY

The study does not involve testing or providing any care or investigation.

POSSIBLE RISKS, DISCOMFORTS AND INCONVENIENCES

There are no risks, discomforts or any inconveniences associated with this research study.

POTENTIAL BENEFlTS

If you participate in this study you may reasonably expect to benefit from the trial by knowing more about the concept and benefits of Birth Companion during labour and delivery, which is an important component of the national Labour Room Quality Improvement Initiative.

In addition, your participation may contribute to the knowledge about the gaps in policy and guidelines of the national Labour Room Quality Improvement Initiative.

SUBJECT’S RIGHTS

Your participation in this study is entirely voluntary. Your questions will be answered clearly and to your satisfaction.

In the event of any new information becoming available that may be relevant to your willingness to continue in this study, you or your legal representative will be informed in a timely manner by the Principal Investigator or his/her representative.

By signing and participating in the trial, you do not waive any of your legal rights to revoke your consent and withdraw from the trial at any time.

CONFIDENTIALITY OF STUDY AND MEDICAL RECORDS

Information collected for this study will be kept confidential. Your records, to the extent of the applicable laws and regulations, will not be made publicly available. Only your lnvestigator(s) will have access to the confidential information being collected.

By signing the informed Consent Form attached, you or your legal representative is authorizing such access to your study records.

Data collected and entered into the Case Report Forms are the property of MAMC. In the event of any publication regarding this study, your identity will remain confidential.

COSTS OF PARTICIPATION

Other than 10 minutes of your time, there are no costs involved in your participation in this study. You will not receive any compensation for participating in this study.

RESEARCH RELATED lNJURY AND COMPENSATION

Since the study does not involve any intervention or test, there is no question of any research related injury of compensation.

WHO TO CONTACT IF YOU HAVE QUESTIONS

If you have questions about this research study and your rights you may contact the Principal Investigator – ……….. E-mail …. or .. Department of Obs. & Gynae, MAMC.

Protocol Title:

## Annexure-2

CONSENT BY RESEARCH SUBJECT

Details of Research Study

Principal Investigator:

…………………. MBBS Student &

…………………. Department of Obs. & Gynae, MAMC.

Subject’s Particulars

Name:

Sex: Femaie/Male

Part I — to be filled by participant

I (Name of Participant) agree / do not agree to participate in the research study as described and on the terms set out in the Participant Information Sheet. The nature of my participation in the proposed research study has been explained to me in

(Language dialect) by …………………….. (PI).

I have fully discussed and understood the purpose of this study. I have been given the Participant Information Sheet and the opportunity to ask questions about this study and have received satisfactory answers and information.

I understand that my participation is voluntary and that I am free to withdraw at any time, without giving any reasons and without any medical care being affected.

I also give permission for information in my questionnaire to be used for research. In any event of publication, I understand that this information will not bear my name or other identifiers and that due care will be taken to preserve the confidentiality of this information.

(Signature of Participant) Date of Signing

Part IV— Investigator’s Statement

l, the undersigned, certify to the best of my knowledge that the nature and purpose of study was fully explained and clearly understood before the study participant’s signing this informed consent form.

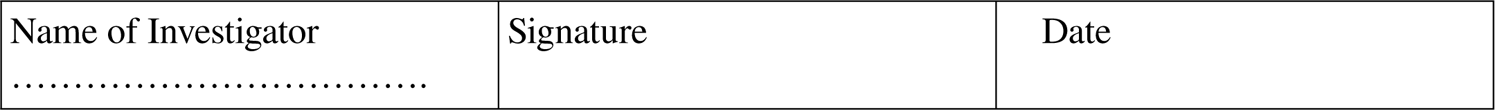

## Annexure-3 Questionnaire for Assessing Awareness regarding, barriers to and suggestions for implementation of Birth Companion in labour and delivery

*Required

1. Name

2. Age

3. Gender *

- Male
- Female

4. Position in Department of Obstetrics and Gynaecology, MAMC / Lok Nayak Hospital:

- Consultant
- Resident
- PG
- Sr. Nurse
- Staff Nurse

4. Are you aware of the concept of Birth Companion?

- Fully Aware
- Somewhat Aware
- Not Aware

5. Are you aware that the WHO recommends every woman to be accompanied by a companion of her choice during labour?

- Fully Aware
- Somewhat Aware
- Not aware

6. Are you aware that the Government of India has recently advocated the presence of a birth companion in all government hospitals?

- Fully Aware
- Somewhat Aware
- Not Aware

7. Who in your opinion can qualify to be a birth companion ?

- Husband / Spouse
- Mother
- Sister
- Mother-in-law
- Children
- Friend
- Paid Daula
- Nurse
- Doctor

8.1 Pre-requisites for a birth companion-should be female relative.

- True
- False

8.2 She should have gone through process of labour ?

- True
- False

8.3 Should not suffer from any communicable disease

- True
- False

8.4 She should wear clean clothes

- True
- False

8.5 She should stay with the pregnant woman throught the process of labour

- True
- False

8.6 She should interfere with the work of hospital staff

- True
- False

8.7 She should attend to other women in the labour room

- True
- False

9. In your opinion, is presence of a birth companion during labour beneficial ?

- Highly beneficial
- Somewhat beneficial
- Not beneficial

10.1 If yes, what in your opinion are the benefits of a birth companionshorter duration of labour

- True
- False

10.2 Reduced need for analgesia.

- True
- False

10.3 Increased spontaneous vaginal births.

- True
- False

10.4 Reduced need for instrumental delivery

- True
- False

10.5 Reduced incidence of unnecessary cessarian sections

- True
- False

10.6 Humanization of labour

- True
- False

10.7 Reduced workload for hospital staff.

- True
- False

10.8 Avoid frequent vaginal examination

- True
- False

10.9 Reduced intrapartum bleeding

- True
- False

10.10 Emotional support

- True
- False

10.11 Comfort measures – soothing touch, message

- True
- False

10.12 Helping the woman to advocate her wish to others

- True
- False

10.13 Delivering in birth position of choice

- True
- False

10.14 Boosting woman’s confidence

- True
- False

10.15 Spiritual support

- True
- False

10.16 Increased satisfaction by women

- True
- False

10.17 Increased use of partograph

- True
- False

10.18 Early initiation of breastfeeding

- True
- False

10.19 Higher newborn Apgar score

- True
- False

10.20 Reduced post-partum depression

- True
- False

11. Will the concept of birth companion be beneficial in dealing with high risk pregnancies ?

- Highly beneficial
- Somewhat beneficial
- Not beneficial

12. Should this concept be introduced in a tertiary care institution like MAMC & Lok Nayak Hospital?

- Strongly agree
- Agree
- Neither agree nor disagree
- Disagree
- Strongly disagree

13.1.1 What according to you are the barriers to implementation of the concept of birth companion in this institution? (tick as appropriate) At the Institution Level: Hospital policy

- Strongly agree
- Agree
- Neither agree not disagree
- Disagree
- Strongly disagree

13.1.2 What according to you are the barriers to implementation of the concept of birth companion in this institution? At Institution level: Overcrowding in Labour room

- Strongly agree
- Agree
- Neither agree not disagree
- Disagree
- Strongly disagree

13.1.3: Lack of privacy for the woman (no dividers or curtains)

- Strongly agree
- Agree
- Neither agree not disagree
- Disagree
- Strongly disagree

13.1.4 Privacy concerns for other women, especially in presence of male companion

- Strongly agree
- Agree
- Neither agree not disagree
- Disagree
- Strongly disagree

13.1.5 Risk of infection transmission

- Strongly agree
- Agree
- Neither agree not disagree
- Disagree
- Strongly disagree

13.1.6 Asepsis in OT will be compromised

- Strongly agree
- Agree
- Neither agree not disagree
- Disagree
- Strongly disagree

13.1.7 Women would not cooperate with hospital staff in presence of birth companion

- Strongly agree
- Agree
- Neither agree not disagree
- Disagree
- Strongly disagree

13.1.8 Use of traditional medicines by birth companion

- Strongly agree
- Agree
- Neither agree not disagree
- Disagree
- Strongly disagree

13.1.9 Birth companion may interfere with clinical decision making

- Strongly agree
- Agree
- Neither agree not disagree
- Disagree
- Strongly disagree

13.1.10 A lounge / waiting area will be needed for the birth companion to take short breaks

- Strongly agree
- Agree
- Neither agree not disagree
- Disagree
- Strongly disagree

13.1.11 The birth companion may not be continuously present during labour

- Strongly agree
- Agree
- Neither agree not disagree
- Disagree
- Strongly disagree

13.1.12 Birth companion will not be able to see the sight of blood

- Strongly agree
- Agree
- Neither agree not disagree
- Disagree
- Strongly disagree

13.1.13 Woman’s health information should be kept confidential

- Strongly agree
- Agree
- Neither agree not disagree
- Disagree
- Strongly disagree

13.1.14 Presence of birth companion is unprofessional

- Strongly agree
- Agree
- Neither agree not disagree
- Disagree
- Strongly disagree

13.2.1 For the patient: Embarrasment of delivering before someone else

- Strongly agree
- Agree
- Neither agree not disagree
- Disagree
- Strongly disagree

13.2.2 Not socially acceptable (stigma / customs)

- Strongly agree
- Agree
- Neither agree not disagree
- Disagree
- Strongly disagree

13.2.3 Birth companion may gossip in the community

- Strongly agree
- Agree
- Neither agree not disagree
- Disagree
- Strongly disagree

13.2.4 Cost of transportation of birth companion

- Strongly agree
- Agree
- Neither agree not disagree
- Disagree
- Strongly disagree

13.2.5 Economic loss if birth companion is an earning member and has to take break from work

- Strongly agree
- Agree
- Neither agree not disagree
- Disagree
- Strongly disagree

13.2.6 Hiring a doula is expensive

- Strongly agree
- Agree
- Neither agree not disagree
- Disagree
- Strongly disagree

13.2.7 No trustworthy person to accompany her as a birth companion

- Strongly agree
- Agree
- Neither agree not disagree
- Disagree
- Strongly disagree

14.1 How can these barriers be overcome-Increased awareness among the hospital staff about the benefits of birth companion

- Strongly agree
- Agree
- Neither agree not disagree
- Disagree
- Strongly disagree

14.2 creating physical partition to ensure privacy

- Strongly agree
- Agree
- Neither agree not disagree
- Disagree
- Strongly disagree

14.3 providing incentives to women delivering in the presence of birth companion

- Strongly agree
- Agree
- Neither agree not disagree
- Disagree
- Strongly disagree

14.4 prior training of birth companion on their role

- Strongly agree
- Agree
- Neither agree not disagree
- Disagree
- Strongly disagree

15.1 Provide funding to hospitals to upgrade labour rooms

- Strongly agree
- Agree
- Neither agree not disagree
- Disagree
- Strongly disagree

15.2 Incentivize hospitals that allow birth companion

- Strongly agree
- Agree
- Neither agree not disagree
- Disagree
- Strongly disagree

15.3 formulate guidelines for instructing birth companion

- Strongly agree
- Agree
- Neither agree not disagree
- Disagree
- Strongly disagree

